# TerraFlow, A New High Parameter Data Analysis Tool, Reveals Systemic T-cell Exhaustion and Dysfunctional Cytokine Production in Classical Hodgkin Lymphoma

**DOI:** 10.1101/2021.09.10.21263388

**Authors:** Daniel Freeman, Catherine Diefenbach, Linda Lam, Tri Le, Jason Alexandre, Bruce Raphael, Michael Grossbard, David Kaminetzky, Jia Ruan, Pratip K. Chattopadhyay

**Author notes:** Authors contributed equally (Daniel Freeman, Catherine Diefenbach).

## Abstract

The incredible variety of proteins associated with immune responses presents a major challenge in immune monitoring. When combinations of these proteins are measured, cell types that influence disease can be precisely identified. Here, we introduce TerraFlow, a novel data analysis tool that performs an exhaustive search of disease-associated cell populations from high-parameter flow cytometry experiments. Using a newly generated dataset, from 24-color immune checkpoint-focused and 18-color immune function-focused experiments, we apply TerraFlow to classical Hodgkin lymphoma (cHL), where systemic T-cell immunity has not been investigated in detail. We reveal novel immune perturbations in newly diagnosed cHL, as well as persistent immune perturbations after treatment. Newly diagnosed patients have elevated levels of activated (CD278+), exhausted (e.g., CD366+ and CD152+ phenotypes), and IL17-expressing cells, along with diminished levels of naïve and central memory (CD127+) T-cells and fewer IFNγ+ and TNF+ T-cells. Exhaustion signatures are reduced with treatment, but compared to healthy individuals, treated patients still exhibit more activated (CD278+ phenotypes), exhausted (CD366+), and IL17-expressing cells. Notably, TerraFlow identifies more phenotypic differences between patient groups than FlowSOM and CellCNN, often with better predictive power. Finally, we introduce a new non-gating approach for data analysis that obviates the need for time-consuming and subjective setting of fluorescence thresholds. Our results benchmark TerraFlow against common methods, provide mechanistic support for past reports of immune deficiency in cHL, and provide a roadmap for future immunotherapy and biomarker studies.

## Introduction

Immune responses are coordinated by a myriad of proteins distributed across a wide variety of cell types. The presence or absence of particular cell types may significantly influence the immune response in malignancy. In lymphoma, for example, the immune landscape of the tumor microenvironment (TME) plays a clear role in disease. Patients with classical Hodgkin lymphoma (cHL) possess rare, malignant Hodgkin/Reed Sternberg (HRS) cells that shape their microenvironment to prevent immune surveillance and inhibit cytotoxic immune responses^1-5^. Various mechanisms underlying this phenomenon have been proposed including the secretion of inhibitory cytokines such as TARC (CCL17), attraction of suppressive T helper 2 (Th2) and regulatory T (Treg) cells to the TME, and differentiation of naïve CD4+ T-cells into forkhead box P3 (FoxP3+) regulatory (Treg) T-cells^6-8^.

Less, however, is known about the systemic immune system of patients with cHL. Specifically, are immune perturbations in cHL global or local, and what influence does TME exert on systemic immunity? Several studies suggest systemic immune dysregulation in early and advanced cHL, as demonstrated by poor responses to recall antigens in delayed-type hypersensitivity testing^9^ and systemic elevations of a variety of secreted immune modulators, including CCL17 (TARC)^8-10^, IL6^11^, IL2R^11^, galectin-1^12^, and soluble CD30^13^. Importantly, these studies did not detail the specific cell subsets that are altered in cHL. Characterization of immune cell subsets in lymphoma is important, because systemic immune dysfunction can influence anti-tumor immunity, treatment responses, or immunity to other diseases or to vaccination. Furthermore, the development of prognostic and treatment related biomarkers is most efficient when candidates are identified from peripheral blood, since this sample type is amenable to routine monitoring. Characterizing the systemic immune landscape of cHL will significantly advance immune biomarker development in cHL.

High-parameter flow cytometry is a particularly established and robust platform for characterizing immunophenotypes but can be limited by bottlenecks in data analysis^14^. Unsupervised methods such as *k*-means and FlowSOM partition cells into clusters based on similar protein expression profiles. Downstream analyses then compare cluster abundance between patient groups. While unsupervised methods are useful for detecting natural groupings, they do not account for heterogeneity within clusters. Moreover, many antibody panels do not resolve biologically distinct subtypes^15^. Recently, supervised methods have begun to couple cell representation and disease association into one iterative process. For example, CellCNN uses a convolutional neural network to automatically learn the molecular features of disease-associated cell types. Because supervised models are directly optimized to predict patient groups, they are more sensitive to rare populations than unsupervised methods^16^. However, models may only report a subset of cell types affected by disease. Moreover, extensive downstream analysis is needed to interpret selected populations and develop biomarkers to define them. There remains a need for a method that can clearly define a complete and interpretable set of cell phenotypes associated with disease.

In this paper, we introduce TerraFlow (www.terraflow.app), a novel data analysis tool that performs an exhaustive search of disease-associated cell populations and returns results in a directly interpretable format. Past tools we (and others) developed systematically measured every possible phenotype generated from Boolean combinations of all markers in an antibody panel^15^. Complete enumeration can produce hundreds of significant phenotypes, many of which contain extraneous or overlapping markers. TerraFlow resolves these redundancies by selecting the smallest set of phenotypes that capture major cohort differences. Each phenotype contains the minimum number of markers needed to define the target population; the addition or subtraction of additional markers reduces the phenotype’s association with outcome. Meanwhile, a recursive feature elimination (RFE) module identifies the smallest set of markers that can be used to discriminate patient groups. This information can be used to design large-scale validation or correlates studies on lower-parameter instruments. Because the output of TerraFlow consists of precisely-defined phenotypes, rather than clusters of cells on a plot, cell populations of importance can be easily interpreted, purified by cell sorting, or developed as clinical biomarkers.

TerraFlow also introduces a novel non-gating approach for generating phenotypes. Traditional flow analyses use hand-drawn gates to define populations of interest. While a convenient method for measuring complex phenotypes, manual gates can obscure expression patterns that do not conform to a strict on/off binary. Take, for example, a setting where cells expressing high levels of both IFNγ and TNF are important in disease outcome (like what is observed for polyfunctional T-cells). Whereas traditional gates treat each population as an on/off binary, we introduce a non-gating approach that incorporates the number of expressing cells as well as their relative brightness. Because IFNγ and TNF are each relatively rare, we first transform fluorescent intensities with a sigmoidal function that inflates the value of bright cells over neutral or dim cells. Because the double-positive population may be rare compared to the single-positive populations, we next apply a root product calculation that favors the double-positive population. Finally, we calculate the average cell weight within each patient and compare population abundance between cohorts. We show that the non-gating method approximates Boolean phenotypes while also capturing important variation within the positive and negative regions.

By combining the power of high-parameter flow cytometry with our novel data analysis platform, we investigated whether newly-diagnosed cHL was associated with perturbations in the T-cell compartment and whether these perturbations resolved after treatment. We assayed cellular proteins associated with activation, exhaustion, and suppression of peripheral T-cells. We also studied cytokine expression after *in vitro* polyclonal re-stimulation in the context of cell differentiation, activation, and exhaustion. Our results catalog peripheral immunity in cHL patients with unprecedented depth, revealing previously unappreciated systemic immune abnormalities in cHL patients.

## Methods

### Cell processing and high-parameter flow cytometry

PBMCs were derived from whole blood using density-gradient centrifugation, resuspended at 10 million cells/mL, and cryopreserved at -135° C. Samples were thawed, washed in RPMI (Invitrogen, Carlsbad, CA), and split into two equal aliquots. The first aliquot was stained with a panel of antibodies described in previous work^15,17^ and **Supplemental Table 1** (“Immune Checkpoint Panel”). The second was stimulated with phorbol myristate acetate (PMA) and ionomycin (Sigma, St. Louis, MO) in the presence of Golgi Plug containing Brefeldin A (BD Biosciences, San Jose, CA). After four hours, cells were stained with the flow cytometry antibodies described in **Supplemental Table 2** (“Cytokine Panel”). Samples were immediately analyzed on a Symphony Flow Cytometer (BD Biosciences, San Jose, CA). Flow cytometry staining from a representative patient is shown in **Supplemental Figure 1A**.

### Human subjects

Informed consent was obtained from 44 cHL patients treated at NYU Langone Health and New York-Presbyterian Weill Cornell between 2011 and 2016. Blood samples were drawn before treatment and at a three-month follow up. 25 age-matched, cryopreserved, healthy donor PBMCs were also obtained from STEMCELL Technologies (Cambridge, MA). Each cohort compared in this study was represented by 25-33 individuals (**Supplemental Table 2**). Typical of cHL, patients had a median age of 34.5 and a range of 18-90 years. 52% were male and 48% female. Patients had nodular sclerosing (80%), mixed cellularity (10%), lymphocyte rich (3%), and unspecified (2%) histology. Most had stage II disease (64%) followed by stage III (14%) and IV (21%). Patients with active viral infection or autoimmune disease were excluded. For post-treatment analyses, patients had received ABVD +/-consolidative radiation.

### Non-gating combinatorics to generate phenotypes

Fluorescent intensities are linearized with the logicle method (F), rescaled to a variable range [α, β] and transformed with a sigmoidal function (θ) that inflates the expression of brighter cells. For negative markers, the transformation is reversed to favor dimmer cells. Sample expression (s) was calculated by taking the root mean product of the transformed cell values. Formally, expression of a given combinatoric phenotype was defined as:

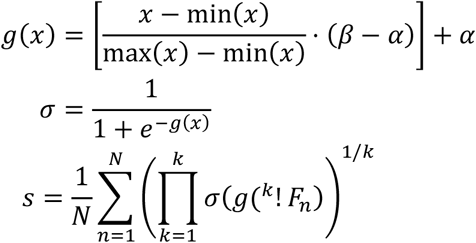

where *F* = Linearized flourescent signal

*K* = Number of proteins

*N* = Number of cells

For example, CD95+CCR7-expression in sample 1 was defined as:

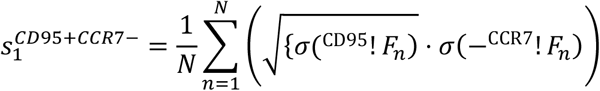

In this study, we used α = −9 and β = 5 for both panels.

### Detection limit

The number of possible combinations increases factorially with the number of proteins. However, fewer cell types are detectable as phenotypes become more complex. TerraFlow excludes phenotypes represented by fewer than 100 cells/sample on average. Samples can have fewer than 100 cells if frequencies follow a non-uniform distribution. For example, a detectable phenotype could be represented by 200 cells/sample in one cohort and completely absent in the other. TerraFlow also excludes gates containing more than 95% of parent events. After filtering out sparse and redundant phenotypes, we found that combinations of 1-5 proteins were sufficient to capture ∼95% of all detectable phenotypes in the Hodgkin’s checkpoint dataset (**Supplemental Figure 1B**). Stratification of healthy and cHL patients did not improve when Elastic Net models were trained on higher-order combinations (alpha=0.1, **Supplemental Figure 1B**). Based on these results, all subsequent results are based on combinations of 1-5 proteins.

### Identification of phenotypes enriched in a patient group

Many canonical immune populations are defined by the combination of proteins they express on their surface. For example, naïve T-cells are defined by CCR7+CD45RA+ while central memory cells are defined by CCR7+CD45RA−. Each additional marker resolves subtypes with deeper levels of granularity. TerraFlow extends this intuition by systematically evaluating every combination of 1 to 5 markers that could be measured within a given panel, generating ∼200,000 phenotypes per dataset. Here, we introduce a novel search algorithm then arranges features into a network that captures gradient changes as markers are successively added, removed, and swapped between related phenotypes. First, linear correlation is calculated between the abundance of each phenotype and patient outcome. Next, phenotypes are arranged into a network by adding edges where nodes differ by the addition or removal of one marker. For example, one path through the network might pass through nodes CCR7+ → CCR7+CD4+ → CCR7+CD4+CD8−. Finally, each node is queried to determine if its correlation is stronger than any adjacent node. These optima are selected to represent unique differences between patient cohorts.

Selected phenotypes are queried again, this time to determine if any markers can be removed without compromising correlation. If a simpler version of a phenotype has a correlation within 97% of the optimal, the extra marker is removed. For example, CD95+CD4+ may have a correlation of 0.90 but CD95+ alone may have a correlation of 0.89. The extra CD4 marker is dropped in favor of the simpler representation. Phenotypes are pruned to convergence, further reducing the total number of selected populations.

Finally, phenotypes are grouped together if they define overlapping cell populations. For example, CD4+ and CD8− both describe helper T-cells. TerraFlow resolves redundant phenotypes by hierarchically clustering populations with Jaccard similarity indices of 50% or greater. Each group is represented as a set of alternative gating strategies or collapsed into the phenotype with the strongest correlation.

Selected phenotypes meet a rigorous, independently verifiable set of criteria. First, each phenotype is represented by an optimal phenotype. Adding additional markers to these phenotypes will not improve association between population abundance and disease status. Conversely, removing any one marker will severely weaken disease association. Second, each phenotype represents a unique cell type. Overlapping cell populations are grouped together, even if defined by distinct molecular features. Finally, each phenotype represents a statistically significant correlate of disease. Together, these criteria allow TerraFlow to return a tractable set of phenotypes without sacrificing important disease information.

### Recursive feature elimination to guide development of simpler antibody panels

While large panels are useful for exploratory purposes, they often contain more markers than are needed to predict the phenotype of interest. TerraFlow uses recursive feature elimination (RFE) to identify the smallest set of markers that allow accurate predictions of patient group. A regularized logistic regression model uses the full set of combinatoric features to predict patient group. Once baseline performance is established, every combination containing the first marker is removed from the feature set and a new model is trained on the remaining phenotypes. The first marker was restored to the feature set and a second marker was removed. The process is repeated for every marker in the panel. At each iteration, the model’s ability to predict sample labels is reevaluated using 10-fold cross-validation and compared to the baseline. The marker whose removal has the least detrimental impact on performance is permanently removed from the feature set. The process repeats until one marker remains. In many cases, performance held constant or even improved until a critical set of markers remained. Removing any of these markers from the panel results in a sharp drop in accuracy. Conversely, restoring any one marker does not significantly improve performance.

### Weighted Lasso maximizes accuracy and interpretability

A custom logistic regression model predicted patient groups during RFE. As in Lasso, an L2 penalty encouraged sparse models by eliminating predictors that were not relevant to the classification task or were highly correlated to each other. Lasso models tend to select complex phenotypes that are overrepresented in the feature set. Here, an additional tiebreaker term penalized predictors based on the number of markers in the phenotype. If two predictors were highly correlated to clinical outcome and each other, the tiebreaker term ensured that the simpler phenotype prevailed.

### Evaluating and confirming model results

Models were evaluated using 10-fold stratified cross-validation. Test set predictions were pooled from each fold before calculating accuracy or area under the curve (AUC). At each fold, we enumerated every combination of 1-5 markers that was detectable in the training set. We then used TerraFlow to identify unique disease-associated cell types. From those, we subset the 20 phenotypes with strongest correlation to patient label or those with an FDR-adjusted P-value smaller than 0.05 (whichever was more stringent). Finally, we trained a ridge logistic regression model to use selected phenotypes to predict patient labels in the test set. Lambda was optimized by performing 10-fold CV within each training set and selecting a value one standard deviation greater than the optimal.

For comparison to existing algorithms, we used the FlowSOM package in R to partition cells into 8 clusters. Fluorescent intensities were logicly-transformed and Z-score normalized. We decreased grid dimensions until the largest cluster contained fewer than 50% of total events. The final model used xdim=5 and ydim=5 for both panels. A simple logistic regression model used cluster frequencies to predict patient labels.

We also used the Python implementation of CellCNN to automatically learn disease-associated cell types. The same preprocessed data was used for FlowSOM and CellCNN analysis. Within each training set, we used nested 3-fold CV to select from the following hyperparameters: maxpool percentages=(0.01, 1.0, 5.0, 20.0, 100.0), nfilters=(3, 4, 5, 6, 7, 8, 9), learning rate=[0.001, 0.01]. We used 3,000 cells per multi-cell input and 200 multi-cell inputs based on previous experiments on PBMCs. To describe selected populations, CellCNN automatically compiled a matrix of filter weights from all runs achieving a validation accuracy above 95%. It then performed hierarchical clustering with a cosine similarity cutoff of 0.4. One representative filter was selected from each cluster to display in the heatmap. We obtained population frequencies by transforming FCS data with each selected filter and measuring the percent of cells with a response greater than 0.

For validation results from the newly-diagnosed comparison to healthy patients, an independent dataset was generated from a new experiment with unique patient samples.

## Results

### TerraFlow: A New High Parameter Data Analysis Pipeline

TerraFlow is a multi-step pipeline for analysis of high parameter single-cell data. It uses an efficient combinatorics approach to enumerate cells expressing every possible combination of up to five markers at a time. **Figure 1** describes each step in the pipeline. First, cell populations are constructed using TerraFlow’s novel “non-gating” approach. Raw fluorescent intensities are transformed with a set of functions that favor bright, multi-positive cells, allowing rare events to affect the overall mean. Expression levels for each marker are automatically translated into the familiar “positive” and “negative” terminology, reflecting relative levels of expression. Thus, cells with high expression are “positive,” while cells with low levels of (or no) expression are called “negative.” TerraFlow can also create Boolean combinations of traditional, user-defined thresholds (i.e., gates, **Figure 1A**). The process is repeated for every possible combination of 1-5 markers, generating ∼200,000 phenotypes per panel.

**Figure 1.**
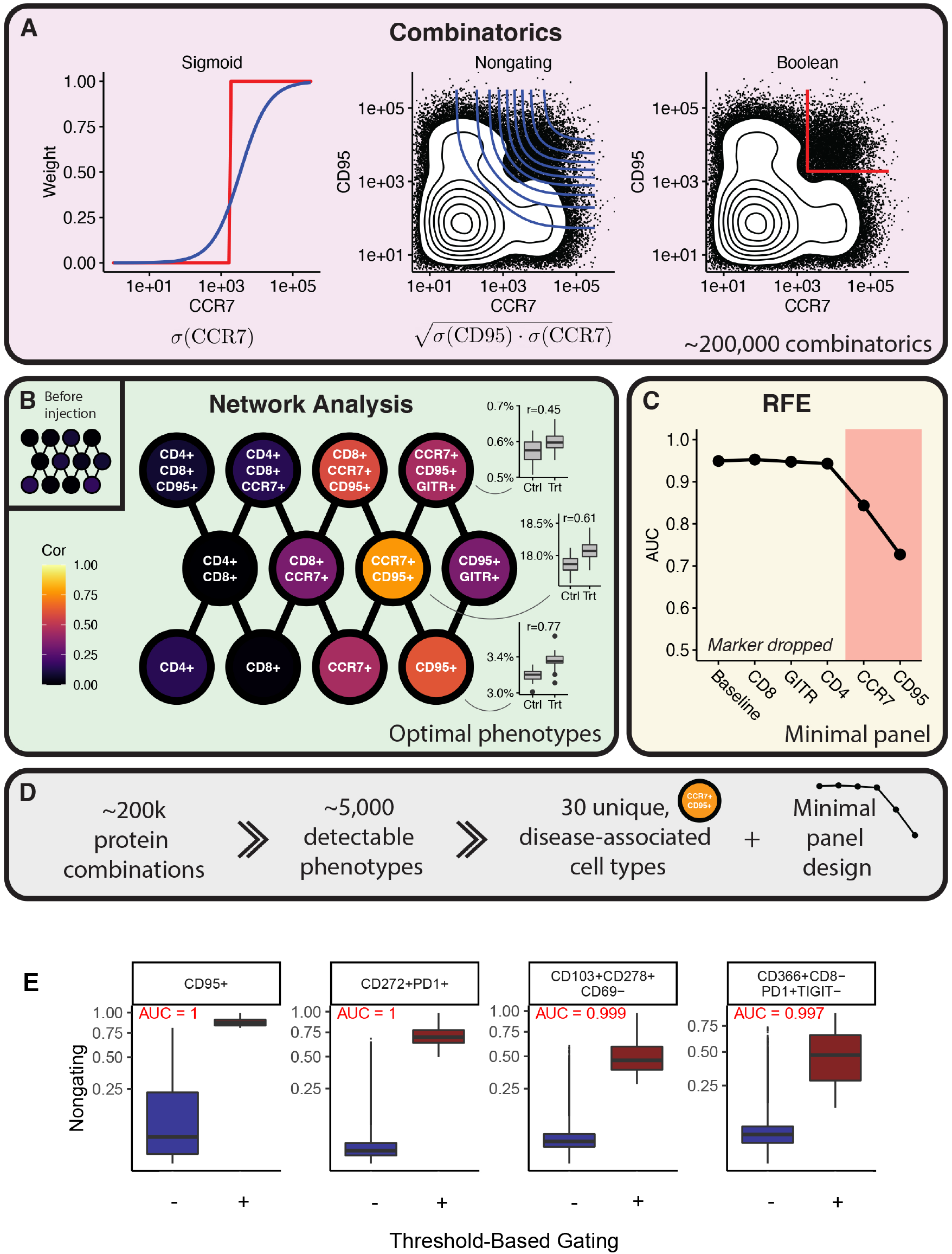
A) TerraFlow introduces a new method for constructing populations by transforming fluorescence signal by a sigmoid function. The non-gating approach weights bright, multi-positive cells heavily. TerraFlow also constructs populations from combinations of gates defined by manual thresholds. B) Populations constructed with either the non-gating or traditional Boolean approaches are tested for their association with study groups, and arranged into a network by adding edges where nodes differ by the addition or removal of one marker. Each node is then queried to determine if its correlation is stronger than any adjacent node. Nodes with the highest correlation to outcome are selected as an optimum representative of a family of cell populations. C) Recursive Feature Elimination (RFE) iteratively tests machine learning models, beginning with a model containing all markers, followed by models that remove one marker at a time. The markers whose removal adversely impacts AUC are those deemed necessary to discriminate the patient groups. D) TerraFlow filters the dataset from ∼200,000 cell populations to identify unique, disease-associated cell types and the minimal set of markers that define the difference between patient groups. E) Comparison between non-gating and threshold-based gating shows that non-gating captures more variation in expression levels from within traditional positive and negative gates.

To identify phenotypes differentially expressed between patient groups, we use a novel form of network analysis (**Figure 1B**). To demonstrate, we simulate a flow cytometry dataset in which there are no differences between two cohorts aside from random noise. We then inject CCR7+CD95+ cells into the treatment cohort. Injection affects the frequency of CCR7+CD95+ but also of related phenotypes such as CCR7+ and CCR7+CD95+GITR+ (boxplots, **Figure 1B**). Rather than report out all three populations as significant, TerraFlow finds the phenotype that best represents cohort differences. First, linear correlation is calculated between population abundance and patient outcome. Next, phenotypes are arranged into a network by adding edges where nodes differ by the addition or removal of one marker. For example, one path through the network might pass through CCR7+ → CCR7+CD95+ → CCR7+CD95+GITR+ (**Figure 1B**). Finally, each node is queried to determine if its correlation is stronger than any adjacent node. These optima are selected to represent unique differences between patient cohorts.

Finally, while large panels are useful for exploratory purposes, they often contain more markers than are needed to predict the clinical outcome of interest. TerraFlow uses recursive feature elimination to identify the smallest set of markers that allow accurate predictions of clinical outcome. A machine learning model uses the entire combinatoric feature set to classify samples (Baseline, **Figure 1C**). The model then iteratively removes the least important marker from the panel and reevaluates performance using 10-fold cross-validation. The process continues until one marker remains. In the simulated dataset, performance remains stable until two markers are left, correctly highlighting the importance of the injected CCR7 and CD95 cells in the simulated dataset (red box, **Figure 1C**). To summarize, the algorithm (**Figure 1D**) constructs ∼200,000 cell populations based on combinations of protein expression, from these about 5,000 are detectable in a typical dataset (see Methods), and network analysis typically identifies around 30 unique, disease associated cell types (as described in the results below). The RFE module also identifies the minimal set of markers that can be used to define the difference between patient groups.

Using data generated from our study of cHL, we compare non-gating and Boolean analyses. For various 1-4 marker phenotypes from the Checkpoint panel (see Methods), cells identified as positive by user-defined threshold-based gating have higher expression values on the non-gating scale (**Figure 1E**). The non-gating approach also captures extensive variation in expression level within the positive and negative regions. Patient-level expression, as defined by the non-gating approach or the threshold-based approach, is highly correlated for phenotypes containing 1-3 markers, and slightly less correlated for higher-order phenotypes (**Supplemental Figure 2A**). Finally, cell populations (i.e., phenotypes) associated with healthy controls or newly-diagnosed-cHL patients have similar associations with outcome, regardless of whether they are defined by the non-gating or threshold-based approaches. These correlations are very high for populations defined by a single marker (1N, **Supplemental Figure 2B**), and good for populations defined by two (2N) or three (3N) markers. Populations defined by more markers show less correlation. Nevertheless, since various features of the algorithm favor simpler phenotypes, the non-gating approach performance is strong, saving the time needed for manual, threshold-based gating.

We use TerraFlow to explore several clinical research questions in the setting of cHL. First, we asked whether measures of T-cell phenotype and function, such as cell activation, exhaustion, and/or cytokine production, are impaired in newly diagnosed cHL patients, compared to healthy controls. Next, we asked whether any differences emerged or persisted after treatment. These analyses compared pre- and post-treatment patients, as well as post-treatment patients and healthy donors. Results are benchmarked against popular methods such as UMAP, FlowSOM, and CellCNN.

### Mapping the topology of T-cell phenotype and function in newly-diagnosed cHL patients

#### Immunophenotypes

We first ask whether systemic T-cell functions such as activation, exhaustion, and suppression were impaired in newly diagnosed cHL patients compared to healthy controls. Our examination begins with TerraFlow’s network analysis (**Figure 1B**). Our non-gating approach evaluated every combination of 1-5 markers that could be formed within the immune checkpoint flow cytometry panel (i.e., Checkpoint dataset), generating approximately 230,000 phenotypes. Of those, approximately 4,800 phenotypes were expressed at detectable levels (see Methods for description of detection threshold). 313 were significantly overexpressed in healthy or newly-diagnosed cHL patients (FDR-adjusted P<0.01). Of those, TerraFlow defined 30 optimal phenotypes. Overlapping populations were further grouped together to produce 27 unique disease-associated cell types. **Figure 2A** describes the top eight phenotypes with the strongest correlation to patient groups (in this case, healthy donors vs. pre-treatment cHL patients). **Figure 2B** defines the expression level of other markers (columns) for 12 immunophenotypes (rows) within a heatmap. The heatmap feature allows investigators to quickly scan expression of other markers, beyond those within the 1-5 parameter phenotypes initially defined by Terraflow. This feature provides more biological insight into each population. For each of the phenotypes, the correlation with outcome is also depicted (right side of the panel); the phenotypes are ordered by the strength and directionality of their correlation.

**Figure 2.**
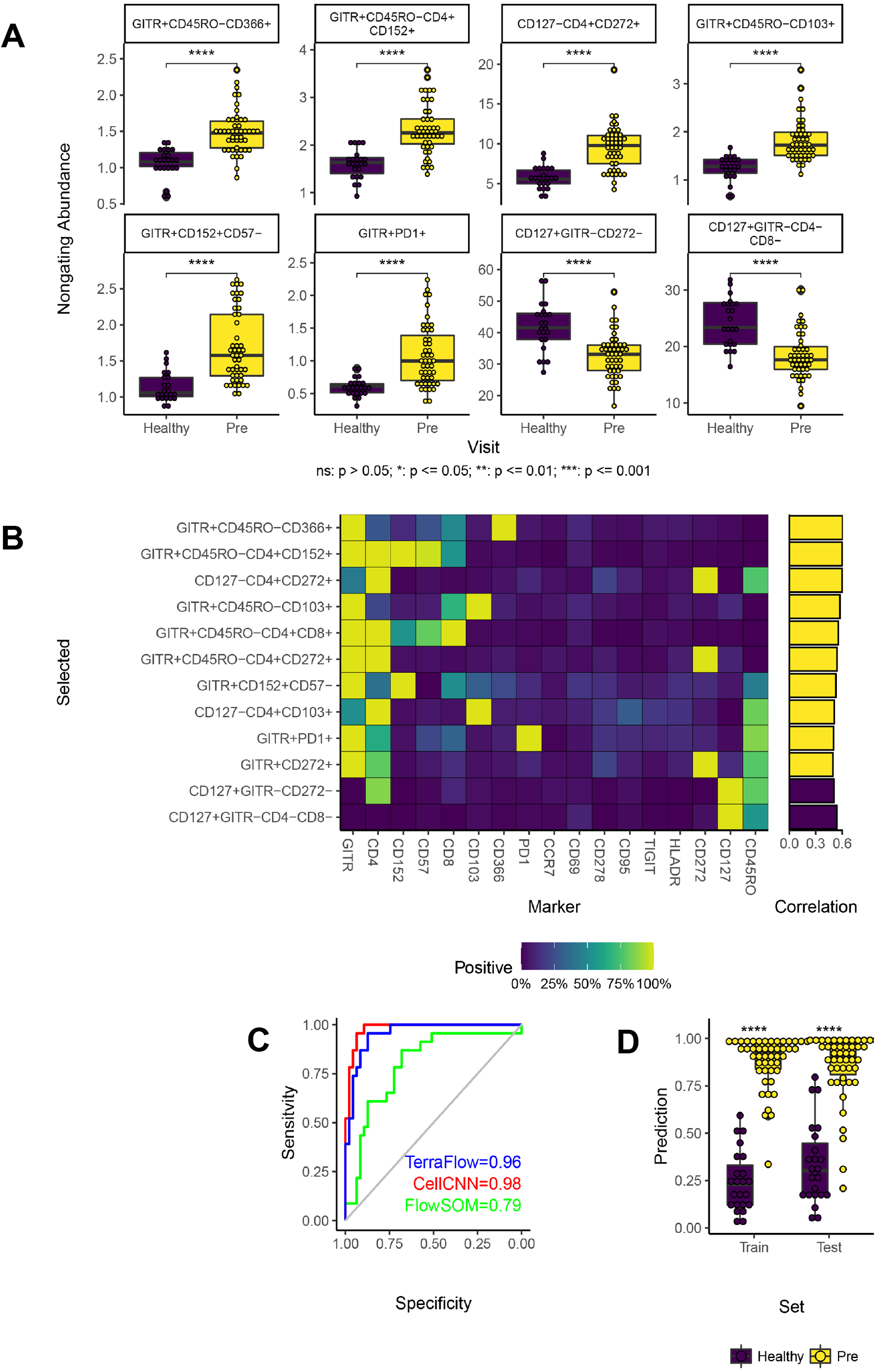
A) Distributions for most statistically significant immunophenotypes across patient groups (healthy vs. newly-diagnosed cHL; Checkpoint Panel). B) Heat map depicting marker frequency (columns) within each phenotype. Adjacent bar graph shows correlation between population frequency and patient group. C) Classification of healthy and newly-diagnosed patients using phenotypes identified by TerraFlow in a ridge logistic regression model; results are compared to CellCNN and FlowSOM. D) Validation of model with training-test set approach.

Models are evaluated using 10-fold cross-validation. TerraFlow achieves excellent separation between healthy and cHL patients, outperforming FlowSOM and approaching CellCNN (AUC=0.96, P<0.001, **Figures 2C and 2D**). Additional validation steps are described later in this manuscript.

Our results reveal that cell populations expressing combinations of GITR, CD366, CD152, CD272, and PD1 are the most enriched in newly diagnosed cHL patients versus healthy individuals (e.g., GITR+CD45RO-CD366+, **Figure 2A**). In contrast, cells expressing CD127 are enriched in healthy individuals (and thus diminished in cHL patients, e.g., CD127+GITR-CD272-, **Figures 2A and 2B**). In sum, these results suggest increased exhaustion (elevated frequencies of GITR+, CD366+, CD152+, and/or PD1+ subsets), activation (CD272+ cells), and differentiation (reduced CD127) of peripheral T-cells in cHL patients.

TerraFlow can also identify the minimal combination of markers that distinguish two study groups through Recursive Feature Elimination (RFE). TerraFlow first trains a regularized logistic regression model to use the full non-gating combinatoric feature set to classify healthy and newly-diagnosed cHL patients. It then iteratively removes the least predictive marker from the panel and reevaluates performance using 10-fold cross-validation. Performance improved as markers were removed from the panel until eight remained. Six markers achieved performance within 95% of the optimal, highlighting the importance of PD1, CD103, CCR7, and GITR (**Figure 3A**). A machine learning model that only includes the RFE-selected markers in combination (**Figure 3B**) distinguishes newly-diagnosed cHL patients from healthy donors in an independent cohort of 20 patients (**Figure 3C**; AUC = 0.97; p<0.001, **Figure 3D**). Cells that are GITR+PD1+ (**Figure 3E**) are significantly higher in cHL patients than healthy controls. Thus, the ensemble of PD1, CD103, CCR7, and GITR represent the simplest set of markers that could be incorporated into a flow cytometry panel to distinguish cHL patients from healthy donors, with GITR+PD1+ cells particularly valuable for identification of newly-diagnosed patients.

**Figure 3.**
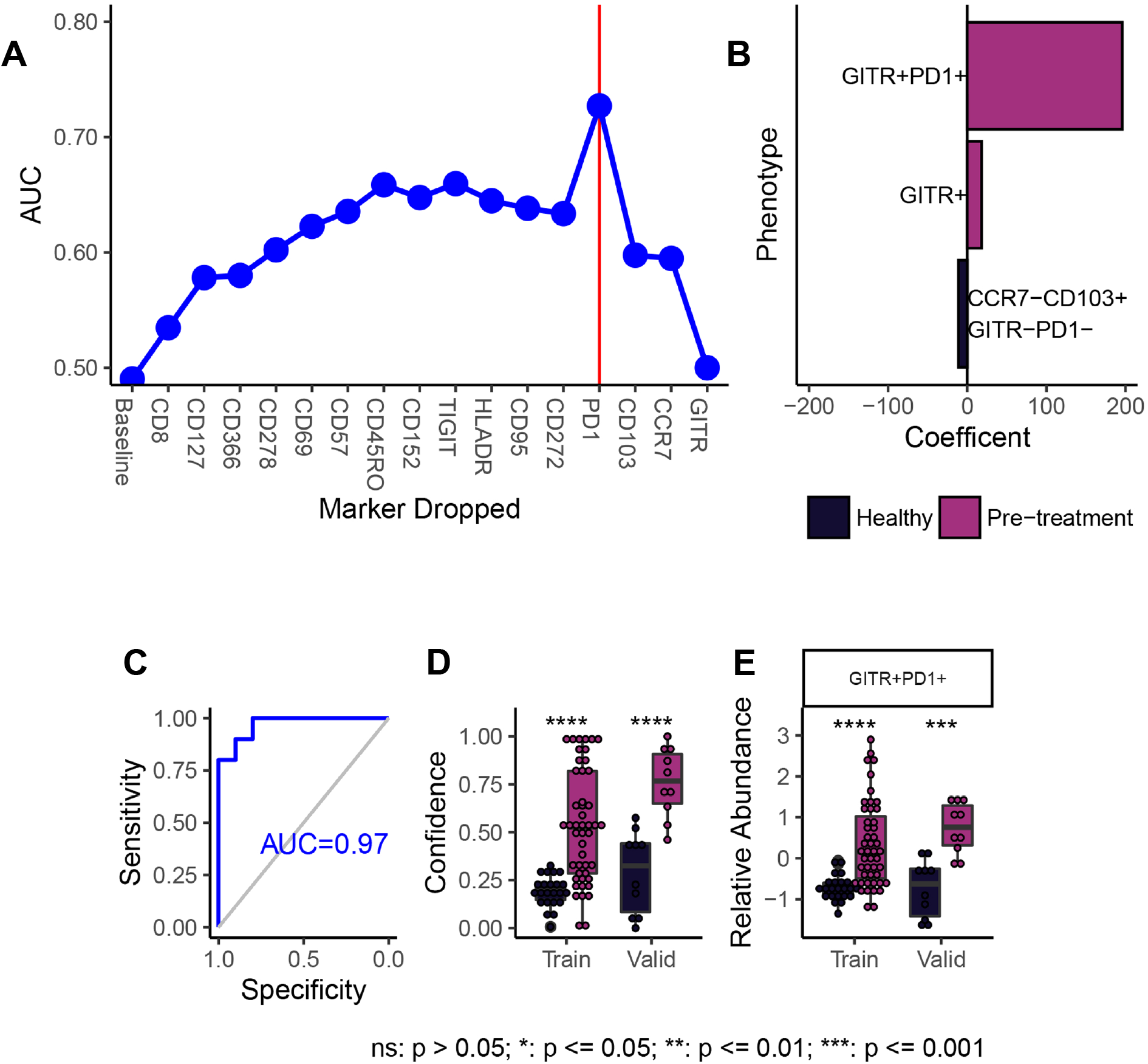
A) RFE identifies PD1, CD103, CCR7, and GITR as a minimal set of markers needed to distinguish healthy donors from newly-diagnosed patients. B) Select phenotypes that contain RFE-selected markers, and their correlation with patient group. C) Machine learning model including only RFE-selected markers distinguishes healthy from newly-diagnosed patients in an independent validation cohort of 20 patients. D) Results from training and validation sets. E) Difference in abundance of GITR+PD1+ cells across patient groups.

#### Traditional Boolean Gating

**Supplemental Figure 3** depicts TerraFlow analysis of cell populations defined by combinations of manually gated thresholds. TerraFlow’s network analysis found that cell populations expressing combinations of GITR, CD152, CD366, CD272, CD278, and HLADR are enriched in newly diagnosed patients (**Supplemental Figures 3A and B**). TerraFlow models trained on Boolean frequencies demonstrate lower performance than models trained on non-gating expression levels (cross-validated AUC=0.88, p<0.0001; **Supplemental Figures 3C and 3D**). Like the non-gating approach, the phenotypes identified in this analysis also suggest that cHL patients exhibit increased exhaustion (GITR+, CD152+, CD366+ phenotypes) and activation (CD278+ and HLADR+ phenotypes).

Recursive feature elimination shows that CD152, CD95, PD1, TIGIT, CCR7, CD8, and GITR are important to define the difference between healthy donors and cHL patients (**Supplemental Figure 3E**). Amongst the cell populations defined by these markers, CD8+CD95+ and CCR7+CD95+ cells have the largest coefficients in the final logistic regression model (**Supplemental Figure 3F**); the complete set of phenotypes formed from these markers can distinguish newly diagnosed patients from healthy donors with high separation (AUC=0.97, p<0.001 for the independent validation study). However, the individual phenotypes with the largest coefficients do not describe statistically significant differences alone (**Supplemental Figure 3F**), suggesting that the full ensemble of RFE-selected markers (CD152, CD95, PD1, TIGIT, CCR7, CD8, and GITR) are required to discriminate patient groups when traditional Boolean gating is used.

#### Immune Function

In the cytokine panel, our non-gating approach generates approximately 1,100 phenotypes significantly overexpressed in healthy or newly-diagnosed cHL patients (FDR-adjusted p<8.5E-6). TerraFlow’s network approach reduces these to 25 unique disease phenotypes (**Figures 4A and B** shows the top phenotypes). TerraFlow distinguishes healthy and newly-diagnosed cHL patients with a cross-validated AUC of 0.82 (not shown), and p<0.001), comparable to FlowSOM (AUC=0.85) but lower than CellCNN (AUC=0.96, not shown). Still, Terraflow defines more disease-associated cell types than either alternative.

**Figure 4.**
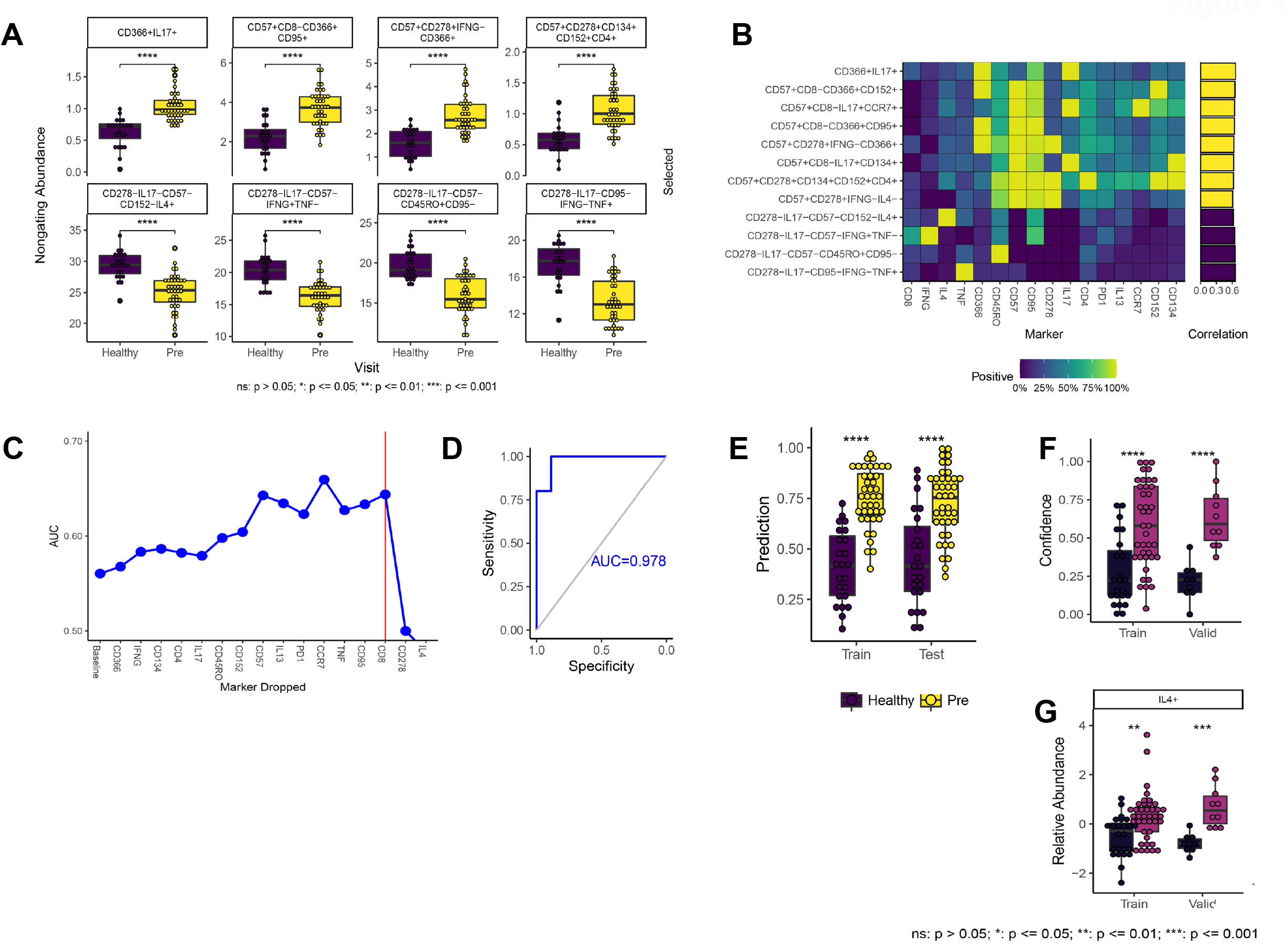
A) Distributions for most statistically significant immune function phenotypes across patient groups (healthy vs. newly-diagnosed cHL; Cytokine Panel). B) Heat map depicting marker frequency (columns) within each phenotype. Adjacent bar graph shows correlation between population frequency and patient group. C) RFE identifies IL4 and CD278 as a minimal set of markers needed to distinguish healthy donors from newly-diagnosed patients. D) Machine learning model including only RFE-selected markers distinguishes healthy from newly-diagnosed patients in an independent validation cohort of 20 patients. E) Results from 10-fold cross validation with training and test datasets. F) Results from training and independent validation cohort of 20 patients. G) Difference in abundance of IL4+ cells across patient groups.

Our results reveal that cell populations expressing combinations of CD152, CD366, CD57, CD95, CD278, CD134 and IL17 are the most enriched in newly diagnosed cHL patients versus healthy individuals. In contrast, cells expressing IFNγ, TNF, and/or IL4 are enriched in healthy individuals (**Figures 4A and 4B**). In sum, these results suggest that peripheral T-cells in cHL patients are exhausted and skewed toward Th17 and Tc17 responses, with a loss of IFNγ, TNF, and IL4-producing cells.

RFE analysis shows that just two markers, CD278 and IL4 (**Figure 4C**), are sufficient to distinguish newly-diagnosed cHL patients and healthy donors (AUC=0.978, p<0.0001 in the independent validation data; **Figure 4D-F**). IL4 distinguishes patients particularly well (**Figure 4G)**. Manual, threshold-based Boolean analysis provided similar results (**Supplemental Figure 4A-D**) but did not identify a reduced set of markers for identification of newly-diagnosed patients (data not shown).

#### Comparison to Common Analysis Approach

We next compared TerraFlow to popular methods such as UMAP, FlowSOM, and CellCNN. UMAP requires visual inspection to identify differences between patient groups, a subjective and time-consuming process. FlowSOM introduces more rigor by automatically clustering cells with similar attributes. FlowSOM results were sensitive to multiple tuning parameters. Furthermore, clusters were contiguous or overlapping in the UMAP projection, reflecting the lack of obvious subtypes in the checkpoint dataset **(Supplemental Figure 4E)**. FlowSOM identified three clusters significantly enriched in newly-diagnosed cHL patients. Of those, only one cluster validated in the follow-up experiment (P<0.05). CellCNN learned populations that were stronger correlates of cHL but only reported two phenotypes (**Supplemental Figure 4F**). Populations are described using mean fluorescent intensities (for FlowSOM) or learned filter weights (for CellCNN, **Supplemental Figure 4G**). However, because populations are defined with complex transformations of the entire panel, it is not clear if a smaller set of markers would have been sufficient to capture the population of interest. Phenotypes could not be validated by manually gating populations in traditional flow cytometry software.

### Changes in T-cell phenotype and function with treatment for cHL

#### Immunophenotype and Function

TerraFlow’s non-gating approach identifies 387 phenotypes that change significantly (FDR-adjusted p<0.05) between the paired comparison of patients before and after their treatment. Amongst these phenotypes, 12 are unique, including those expressing combinations of PD1+ and CD366+ (elevated pre-treatment) and those expressing HLA-DR+, CD95+, or TIGIT+ (elevated post-treatment; **Supplemental Figures 5A and 5B**). During cross-validation, TerraFlow correctly identified the post-treatment sample in 83.3% of individuals, rivaling CellCNN (85.2%) and outperforming FlowSOM (62.3%, **Supplemental Figure 5C;** TerraFlow validation results shown in **Supplemental Figure 5D**). These results suggest that circulating exhausted cells before treatment are replaced by activated (HLADR+) cells, with one exhausted TIGIT+ cell population persisting post-treatment.

RFE analysis (data not shown) could not identify a minimal set of markers from the Checkpoint panel that could distinguish pre- and post-treatment time points. Traditional Boolean analysis, based on investigator-defined thresholds, also could not identify an ensemble of Checkpoint panel phenotypes that distinguished pre- and post-treatment in a machine learning model in the validation dataset (data not shown, mean accuracy = 55%). For the cytokine panel, there was no statistically significant difference observed between pre- and post-treatment, with either the non-gating or traditional Boolean gating approaches (data not shown).

### Do T-cell phenotype and function normalize after treatment for cHL?

#### Immunophenotypes

For the Checkpoint panel, strong differences were observed between cHL patients after treatment and healthy individuals (26 unique phenotypes detected, TerraFlow AUC=0.94, p<0.0001 with 10-fold cross-validation; data not shown). The performance of TerraFlow’s machine learning model rivaled CellCNN (AUC = 0.96) and exceeded FlowSOM (AUC = 0.79). After treatment, cHL patients continue to exhibit higher levels of activated and exhausted cells than healthy donors, including various cell populations expressing CD272, GITR, or CD152 (**Supplemental Figures 6A and 6B**), as defined with our non-gating approach. Notably, cell populations expressing PD1 are lower in patients after treatment than healthy donors, suggesting heterogeneity in checkpoint responses to treatment, and reinforcing the importance of high-parameter, single-cell analysis of multiple markers of exhaustion (**Supplemental Figures 6A and 6B**). RFE of non-gating data did not identify an interpretable minimal set of markers that distinguish post-treatment patients from healthy donors (data not shown).

Traditional, threshold-based comparison of post-treatment patients and healthy donors gave largely similar results, highlighting in addition the elevation of a CD366+ cell phenotype (data not shown). RFE of the threshold-based data showed that the most important markers for describing immunophenotypic differences between post-treatment patients and healthy controls were CD4, CCR7, CD152, and CD57 (**Supplemental Figure 6C**); models built from the phenotypes that include these markers have an AUC of 0.726, with high statistical significance in the test set (p<0.01; **Supplemental Figure 6D**). In particular, CD152+CD57-cells are elevated post-treatment. The overall pattern from both analysis approaches reveals continued exhaustion of cells post-treatment (as evidenced by CD152, and CD57 phenotypes), without normalization to healthy donor levels.

#### Immune Function

TerraFlow revealed 25 disease-associated differences in functional phenotypes between healthy donors and post-treatment cHL patients. Cell types enriched post-treatment included stimulated cells expressing multiple activation and exhaustion markers (CD366, CD95, CD57, CD278, CD152, CD134; **Figure 5A**), as well as cell populations expressing IL17 (e.g., CD366+ IL17+; **Figure 5B**) and IL4 (CD57+ CD4+ CD278+ CD152+ IL4+; **Figure 5B**). In contrast, cell populations expressing TNF were diminished post-treatment (**Figures 5A and 5B**). Similar results were found in the threshold-based analysis (data not shown). In sum, the data suggests that after treatment, cHL patients have increased polarization of cells toward Th2 and Th17 functions, rather than Th1 function, and cells expressing cytokines in post-treatment patients may be more prone to exhaustion than healthy donors. Our model achieved a cross-validated AUC of 0.91 (p<0.0001), rivaling CellCNN (AUC=0.92) and outperforming FlowSOM (AUC=0.83).

**Figure 5.**
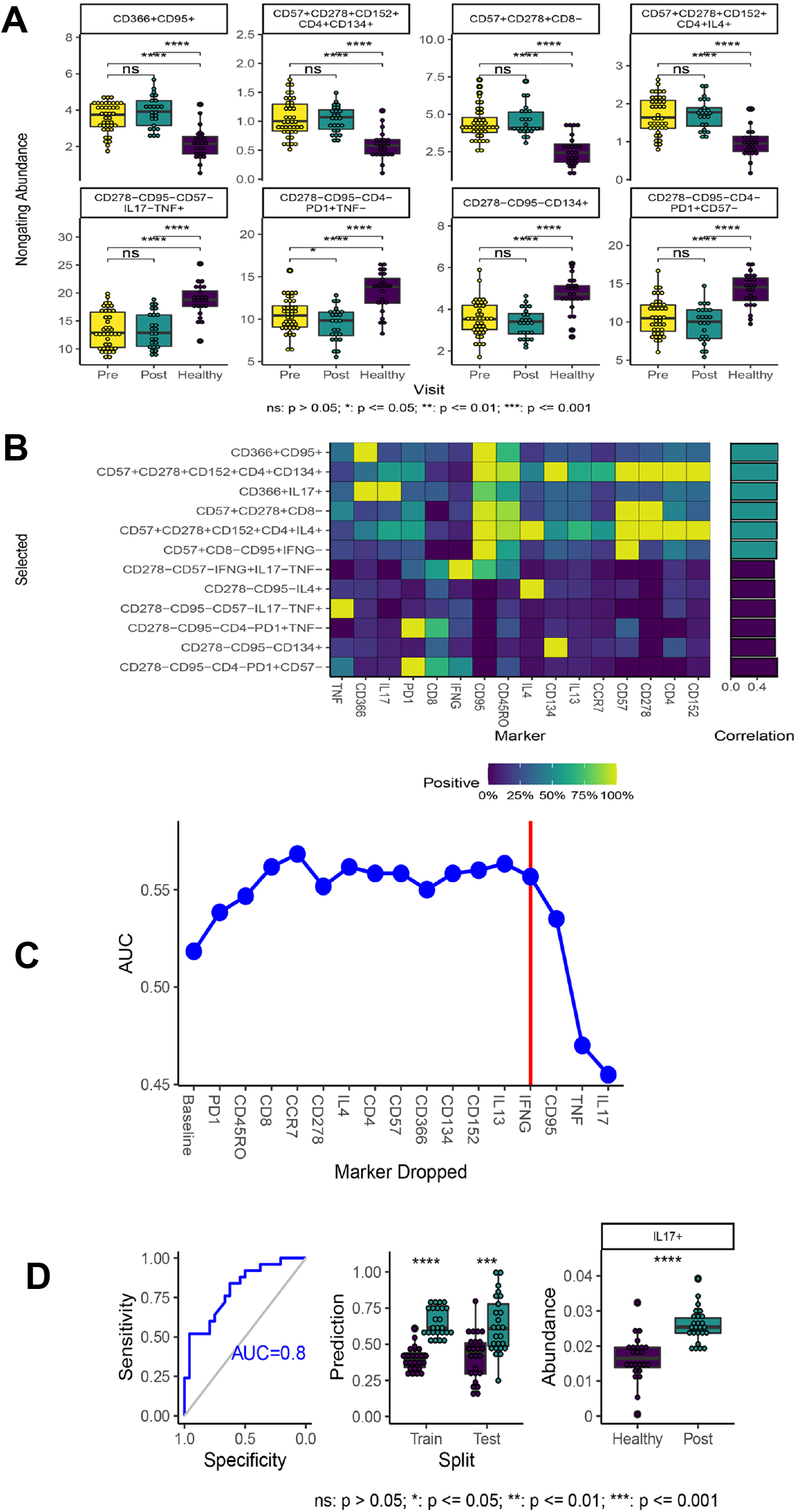
A) Distributions for most statistically significant immune function phenotypes across patient groups (newly-diagnosed vs. post-treatment cHL; Cytokine Panel). B) Heat map depicting marker frequency (columns) within each phenotype. Adjacent bar graph shows correlation between population frequency and patient group. C) RFE identifies CD95, TNF, and IL17 as a minimal set of markers needed to distinguish pre- and post-treatment patients. D) Machine learning model including only RFE-selected markers distinguishes healthy from newly-diagnosed patients in an independent validation cohort of 20 patients. E) Results from 10-fold cross validation with training and test datasets. F) Difference in abundance of IL17+ cells across patient groups.

Recursive feature analysis (**Figure 5C**) of the non-gating data shows that CD95, TNF, and IL17 are the minimal set of markers needed to identify differences between post-treatment patients and healthy donors (cross-validated AUC=0.80; p<0.001; **Figure 5D**). In particular, IL17+ cells are elevated post-treatment compared to healthy donors (p<0.0001, **Figure 5D**). These findings are complementary to, and consistent with, the output of earlier steps in the algorithm: post-treatment patients have reduced Th1 (i.e., IFNγ or TNF) responses, and more exhausted cells primed for apoptosis (CD95+) than healthy donors.

### Selected phenotypes validate to new data

To further demonstrate the concordance between our novel non-gating approach and traditional threshold-based analysis, we selected the 12 phenotypes most significantly over or under-expressed in newly-diagnosed cHL patients compared to healthy donors. We then manually gated cells with the same phenotype in an independent cohort of 20 individuals. Of the 12 phenotypes generated by our non-gating TerraFlow algorithm, 11 validated to new data when measured with traditional, threshold-based gating (p<0.05). Individual populations from the Checkpoint panel (**Figure 6A**) differed strongly between newly-diagnosed cHL patients and healthy donors with a median fold-change of 5.4 and p-value of 0.0044 in the validation set (**Figure 6A**). We then trained a logistic regression model on non-gating expression in the original dataset and applied it to the traditional frequencies in the validation dataset. The model achieved perfect separation between healthy and cHL patients using the 12 selected cell types (p<0.0001), outperforming FlowSOM- and CellCNN-based analyses (**Figure 6A**). Similar results were observed with populations identified from the Cytokine panel analyses, with all three algorithms achieving perfect separation (**Figure 6B**).

**Figure 6.**
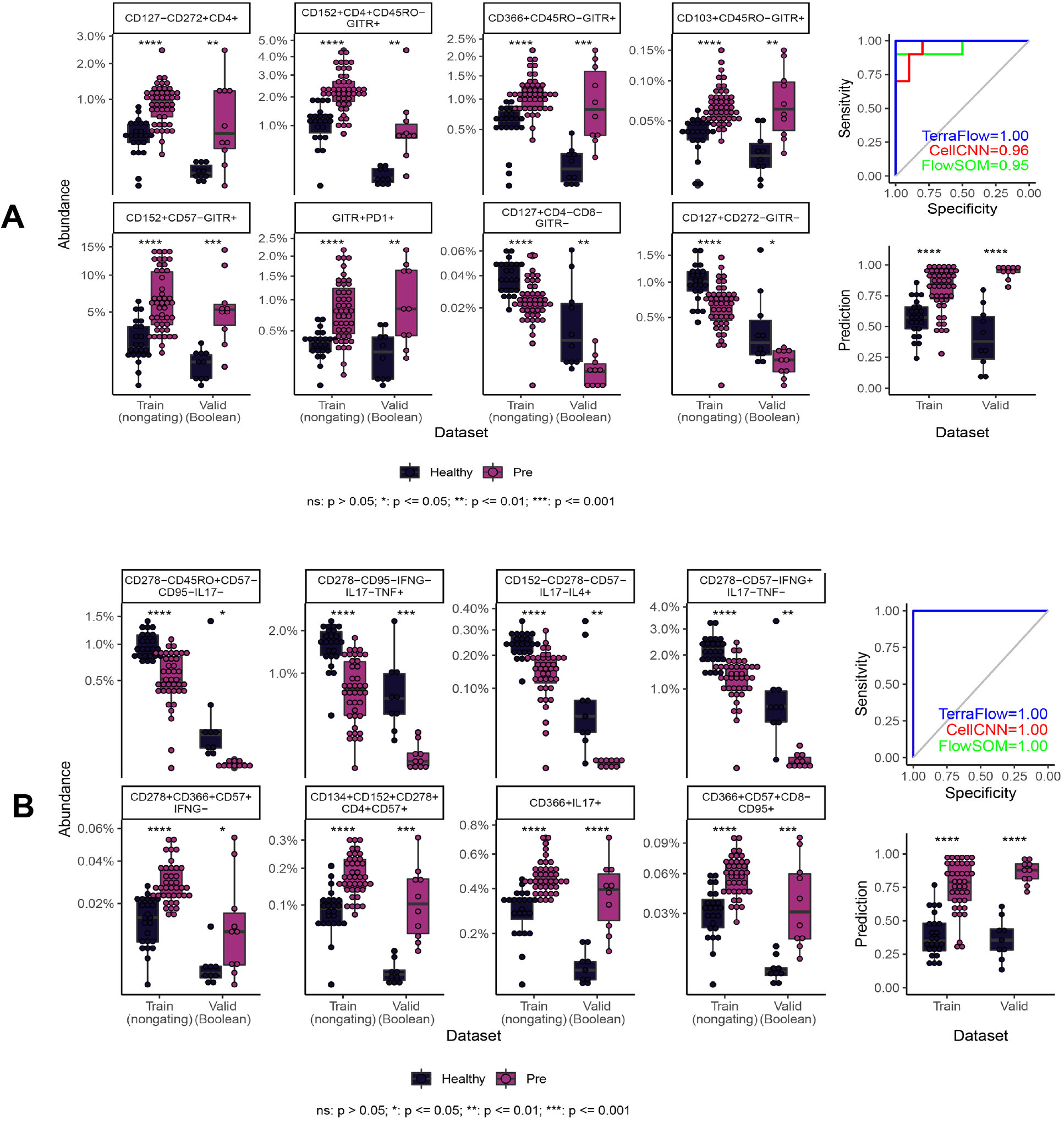
A) Immune checkpoint phenotypes identified in the training dataset with the non-gating approach can be identified by manual gating in the independent validation cohort, and then frequencies can be compared across patient groups to show that results from the non-gating approach are faithfully replicated. B) Immune function profiles, identified with the Cytokine Panel, can also be replicated across non-gating and traditional approaches.

## Discussion

TerraFlow provides several advantages over current data analysis approaches. First, TerraFlow performs an exhaustive search for disease-associated cell types. In the checkpoint panel, FlowSOM identified one cluster that was weakly associated with cHL in a follow-up experiment. CellCNN identified populations that were stronger correlates of cHL but only reported two phenotypes. By contrast, TerraFlow consistently found ten or more unique cell types that were each strongly correlated to cHL and validated to new data.

Second, TerraFlow defines each population with an explicit phenotype. FlowSOM and CellCNN represent populations with complex transformations of the entire panel. By contrast, TerraFlow selects phenotypes that can be described with just one or two markers, only adding three or more if necessary to define the target population. Gating strategies can be directly implemented in traditional flow cytometry software. They can be validated using smaller panels typically used in large typical trials. For deeper characterization, gating strategies can be used to sort populations for downstream experiments such as functional assays or whole-transcriptome sequencing. Whereas traditional methods require extensive downstream interpretation, TerraFlow populations can be directly isolated and developed as putative biomarkers.

Finally, TerraFlow provides superior ease of use. Existing methods require users to anticipate the number of clusters or tune arcane machine learning parameters. By contrast, TerraFlow doesn’t require any input beyond clean FCS data and patient labels. Our non-gating approach even obviates the need for manual thresholds, approximating Boolean gates without using fluorescent cutoffs at all. We show that that the non-gating approach captures expression changes within the target population, increasing overall predictive power. Selected phenotypes easily translate to classical hand-drawn gates.

Our study of patients with cHL provides a rich and finely detailed analysis of the immunophenotypes and functional features of T-cells before and after treatment. Many cell types expressing markers of T-cell exhaustion and activation are elevated in newly-diagnosed (untreated) patients, revealing extensive, systemic alterations in T-cell subset representation in patients. These alterations include changes in the polarization of function in T-cell subsets, as cells are more likely to be TH17 and TC17 cells than TH1 cells in untreated patients (compared to healthy donors). The loss of IFNγ+ cells in cHL patients may release the brakes on IL17-responses. Our results offer potential mechanistic explanations for immune dysfunction in cHL patients. Our results also suggest that other immune checkpoint targets beyond PD1 may be valuable in cHL treatment, such as CD152 (CTLA), CD366 (TIM3), CD278 (ICOS), CD272 (BTLA), TIGIT, GITR, or cell surface CCR4 and CCR6 (to target TH17 cells). Interestingly, around three months post-treatment, patients still exhibit an altered T-cell immune checkpoint and functional landscape, even though the levels of PD1-expressing cells have normalized; future studies will test whether particular alterations in immune checkpoints are associated with cHL relapse.

## Data Availability

Data is not publicly available yet.

## Acknowledgements

The authors would like to thank Suraj Saksena for coordinating the BD Bioscieces/Precision Immunology Laboratory visiting scientist program under which TL began these experiments. We would also like to thank additional investigators who contributed patients to this study: Tibor Moscovits and Kenneth Hymes (NYU Langone Health), as well as Peter Martin and John Leonard (Weill Cornell). We also acknowledge funding support from the Feinberg family, and the American Cancer Society MRSG (CD).

## Author Contributions

DF performed experiments, developed TerraFlow, analyzed data, and wrote manuscript. CD conceived of and designed studies, led the clinical study and recruited patients, analyzed and interpreted data, and wrote manuscript. TL performed experiments. LL organized study and performed experiments. JA organized study and performed experiments. BR, MG, DK, and JR conducted the clinical study and recruited patients. PKC conceived of and designed studies, developed and benchmarked TerraFlow, analyzed and interpreted data, and wrote manuscript.

---

**Supplemental Table 1.**
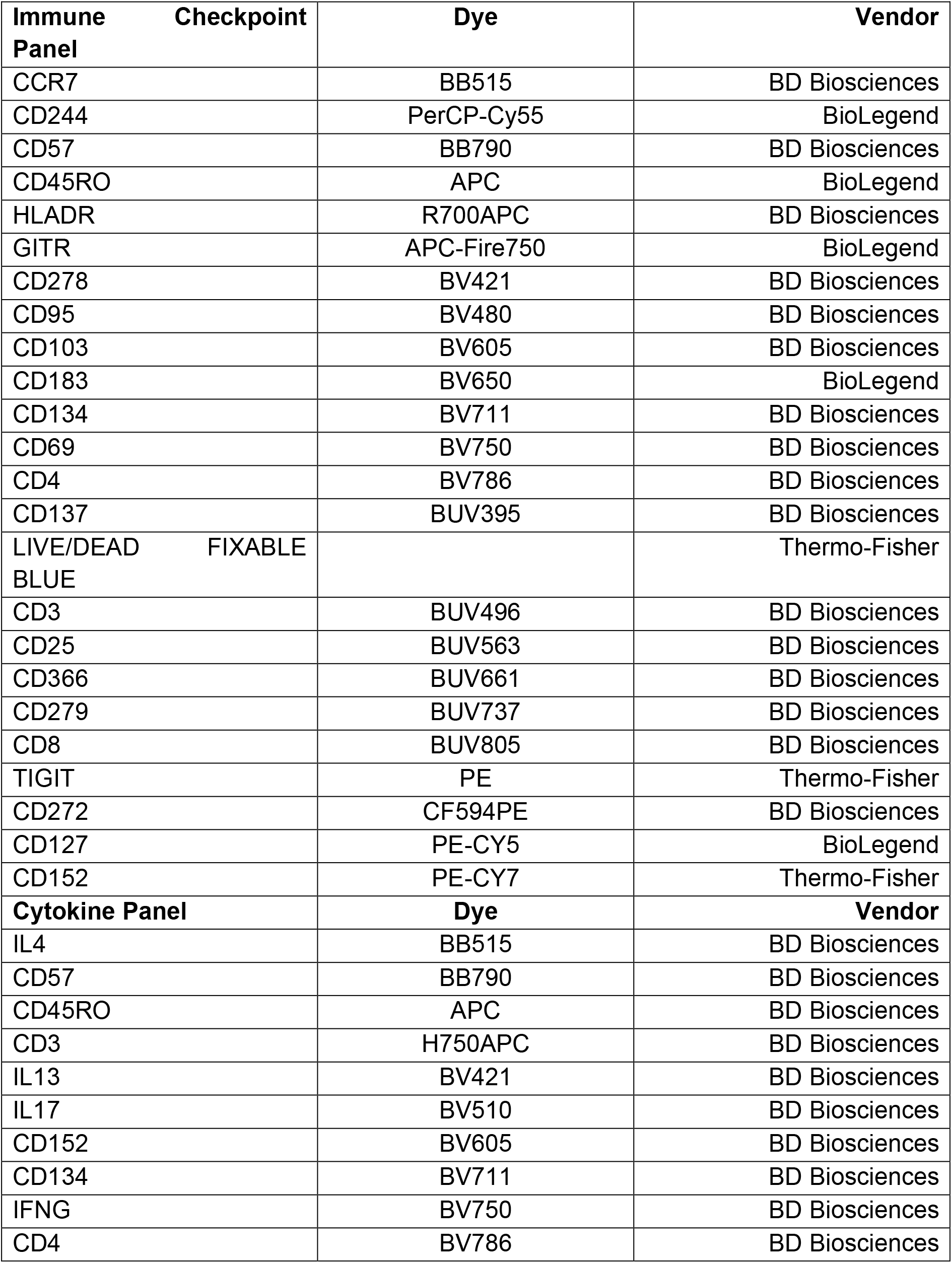
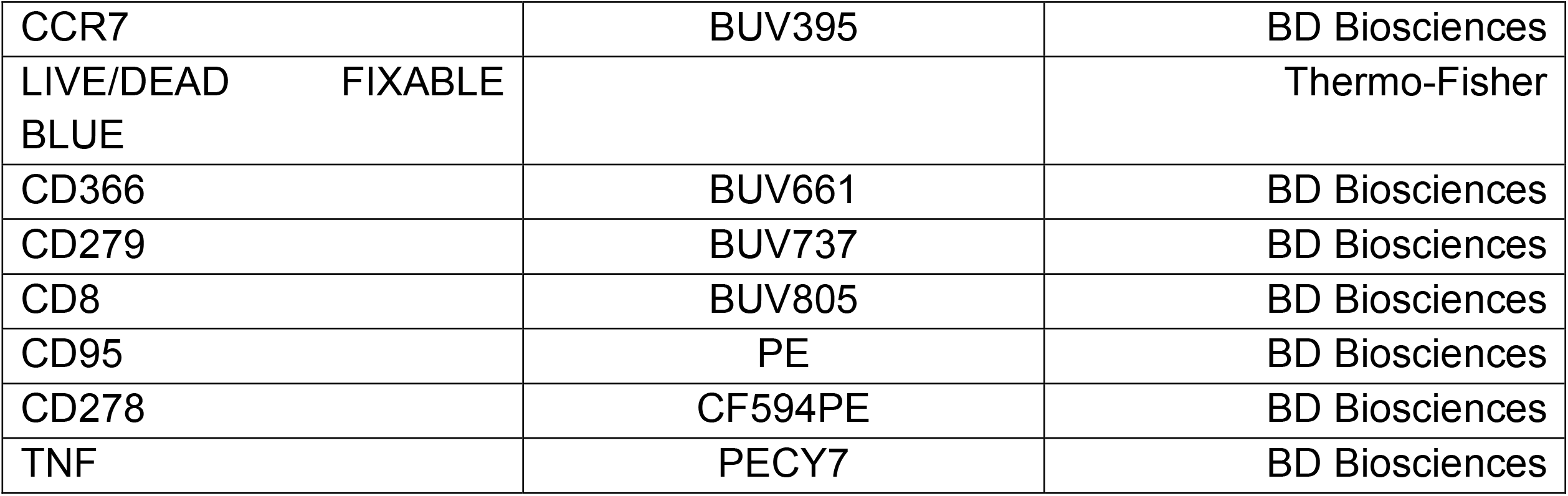

**Supplemental Table 2.**

| **Comparison** | **Panel** | **Group 1** | **Group 2** |
| --- | --- | --- | --- |
| Newly-diagnosed vs. healthy | Checkpoint | 30 cHL | 25 healthy |
|  | Cytokine | 28 cHL | 25 healthy |
| Post-treatment vs. healthy | Checkpoint | 25 cHL | 25 healthy |
|  | Cytokine | 21 cHL | 25 healthy |
| Pre- vs. post-treatment | Checkpoint | 34 pre | 34 post |
|  | Cytokine | 33 pre | 33 post |

**Supplemental Figure 1:**
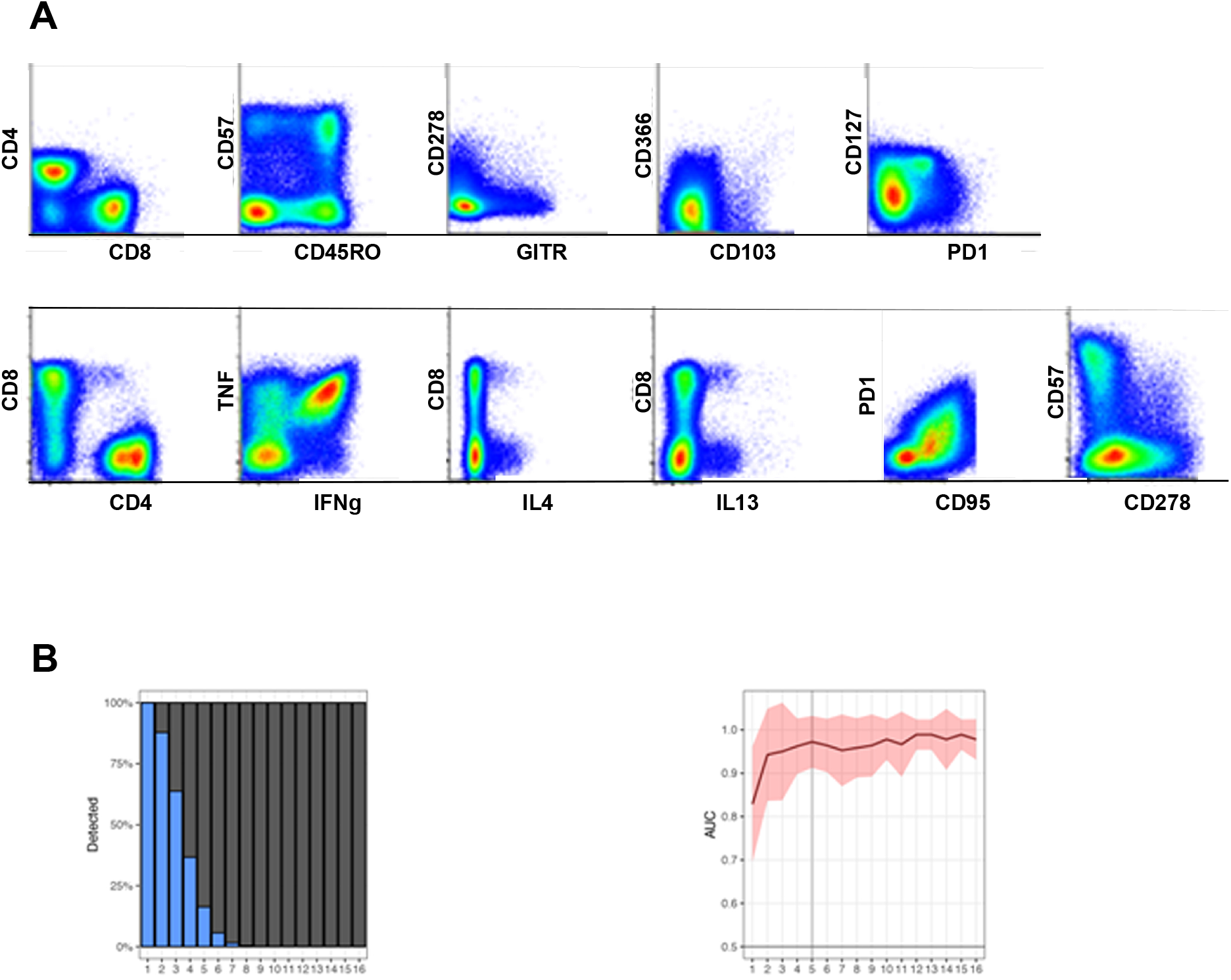
A) Antibody staining from a representative patient. B) Left panel: As the number of parameters in a combinatorial phenotype increase, many of the putative cell populations are undetectable. Beyond five markers, >95% of the theoretical phenotypes produced by exhaustive combinatorics are not represented in the dataset. Right panel: Discrimination of cHL versus healthy patients does not improve as phenotypes contain more than five markers.

**Supplemental Figure 2:**
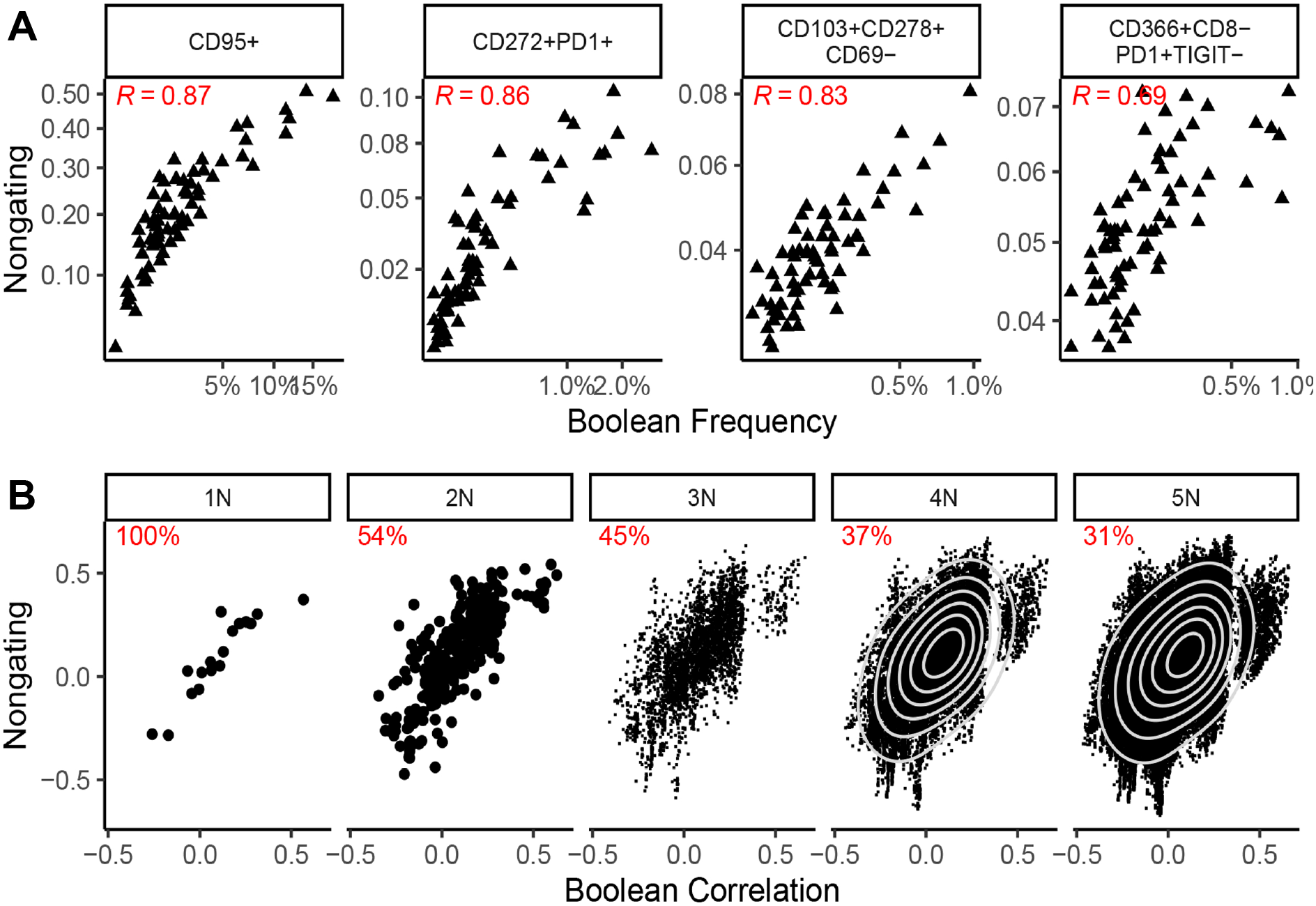
A) Correlation between frequencies of cells identified by non-gating and Boolean approaches, for single, two, three, and four marker phenotypes. B) The association with patient group is correlated for single marker phenotypes regardless of whether non-gating or Boolean gating is used. For more complex phenotypes, defined by 3-5 markers (3N-5N), results are less correlated for the approaches. TerraFlow favors the identification and reporting of simpler phenotypes.

**Supplemental Figure 3.**
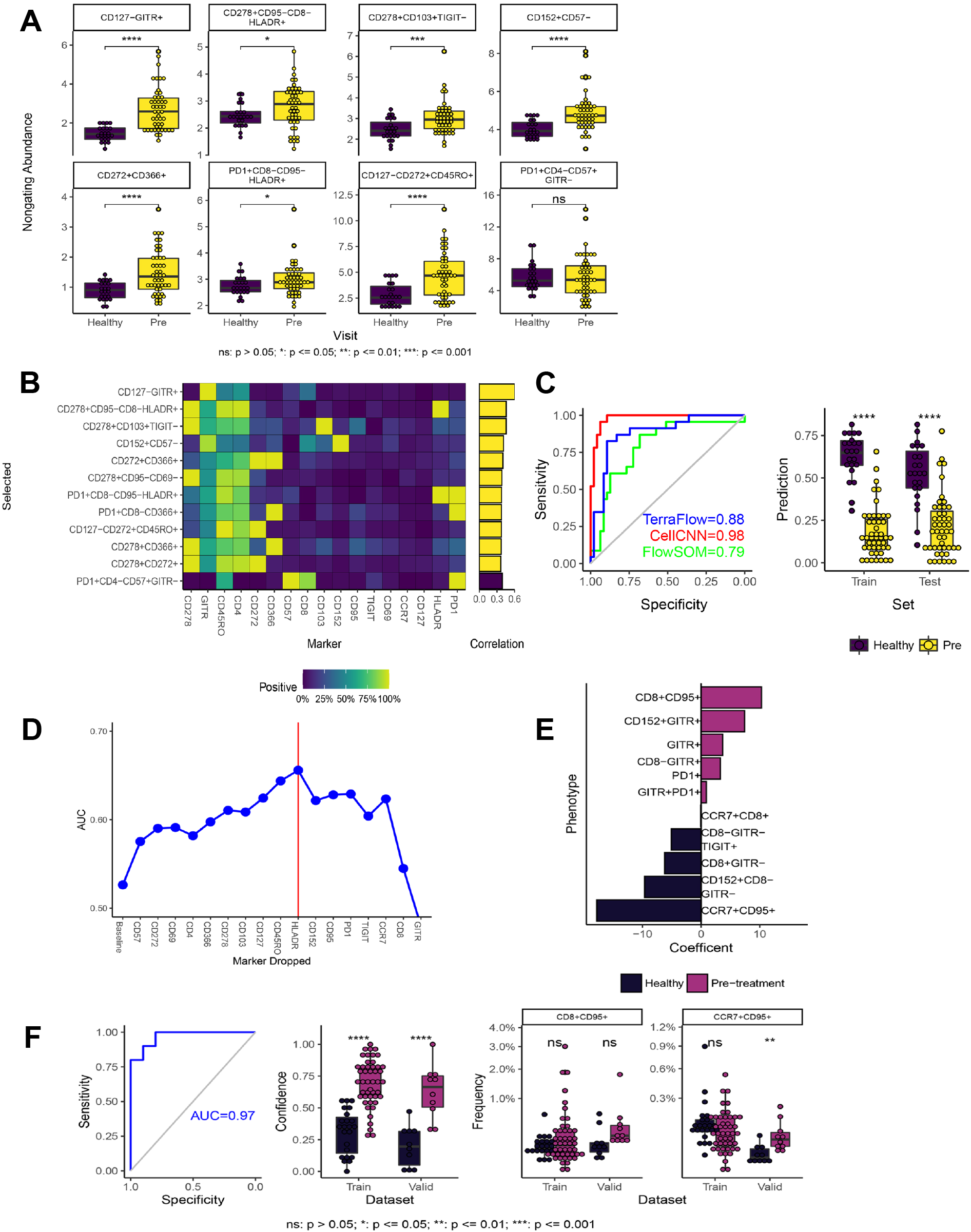
A) Distributions of the most statistically significant phenotypes across patient groups (healthy vs. newly-diagnosed cHL; Checkpoint Panel; traditional Boolean Gating). B) Heat map depicting marker frequency (columns) within each phenotype. Adjacent bar graph shows correlation between population frequency and patient group. C) Classification of healthy and newly-diagnosed patients using phenotypes identified by TerraFlow in a ridge logistic regression model; results are compared to CellCNN and FlowSOM. D) RFE analysis identifies a broad ensemble of markers necessary to identify differences between patient groups. E) Correlation with study group for select phenotypes that contain RFE-selected markers. F) Model, validation, and data for selected RFE-defined phenotypes.

**Supplemental Figure 4.**
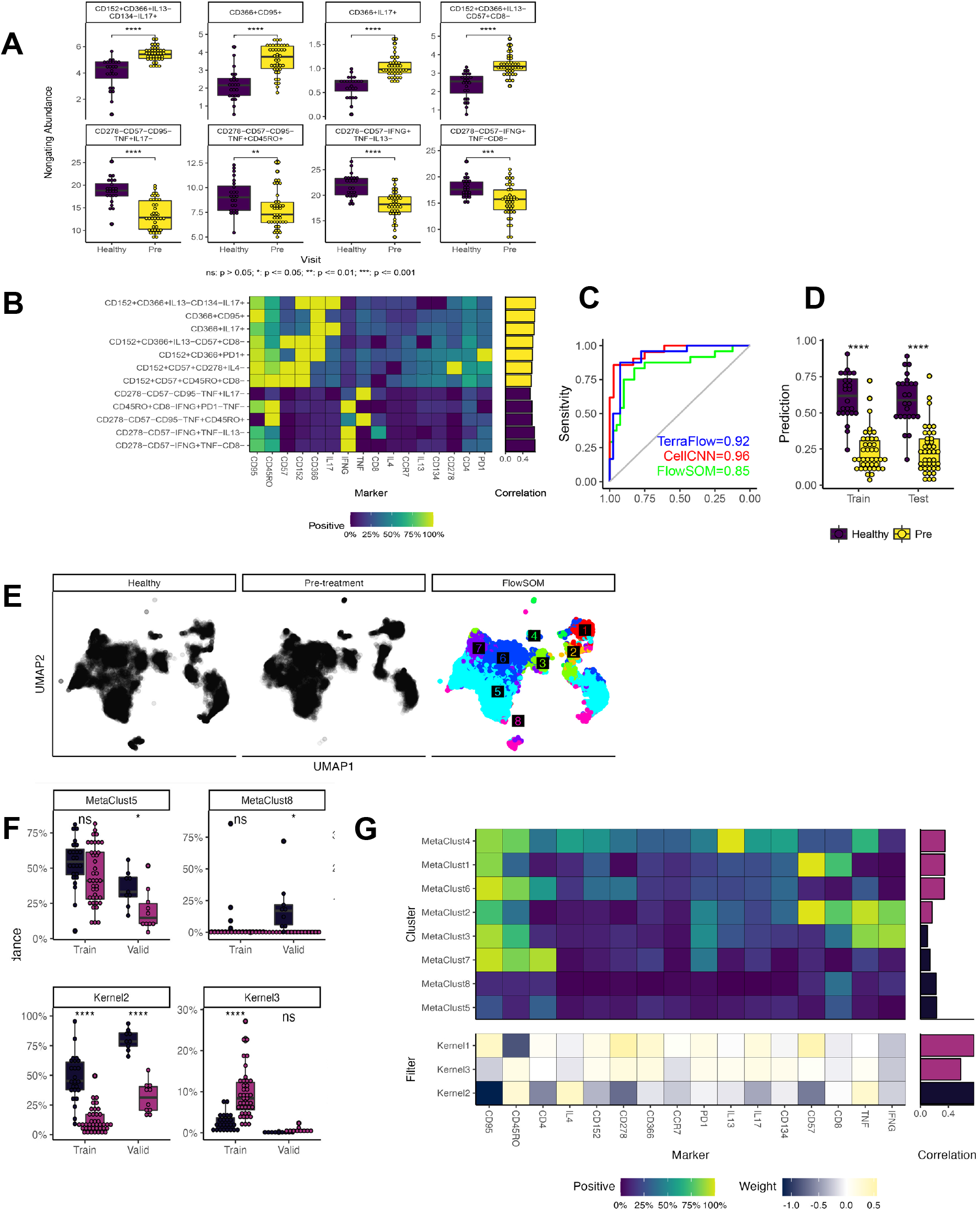
A) Distributions of the most statistically significant phenotypes across patient groups (healthy vs. newly-diagnosed cHL; Cytokine Panel; traditional Boolean Gating). B) Heat map depicting marker frequency (columns) within each phenotype. Adjacent bar graph shows correlation between population frequency and patient group. C) Classification of healthy and newly-diagnosed patients using phenotypes identified by TerraFlow in a ridge logistic regression model; results are compared to CellCNN and FlowSOM. D) Results for 10-fold cross-validation. E) UMAP projections of healthy donor data, pre-treatment patient data, and overlay of FlowSOM-generated clusters. F) Distributions of frequencies for FlowSOM-defined clusters (top) and CellCNN-define populations (bottom) across patient groups. G) Marker expression across clusters (FlowSOM) and Kernels (CellCNN), and correlation with outcome.

**Supplemental Figure 5.**
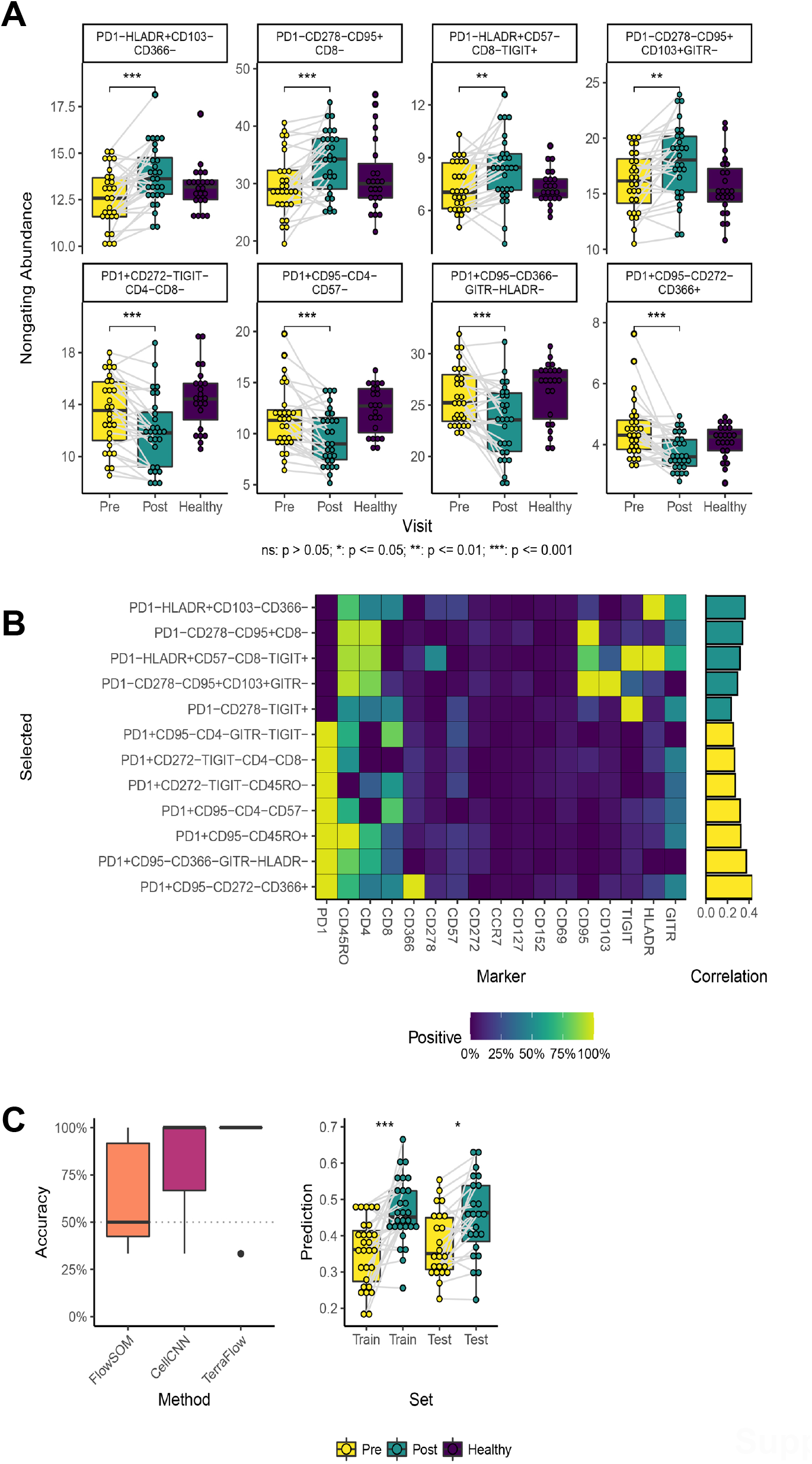
A) Distributions of the most statistically significant phenotypes across patient groups (pre- vs. post-treatment; Checkpoint Panel; non-gating approach). B) Heat map depicting marker frequency (columns) within each phenotype. Adjacent bar graph shows correlation between population frequency and patient group. C) Classification of pre- and post-treatment patients using phenotypes identified by TerraFlow; results are compared to CellCNN and FlowSOM.

**Supplemental Figure 6.**
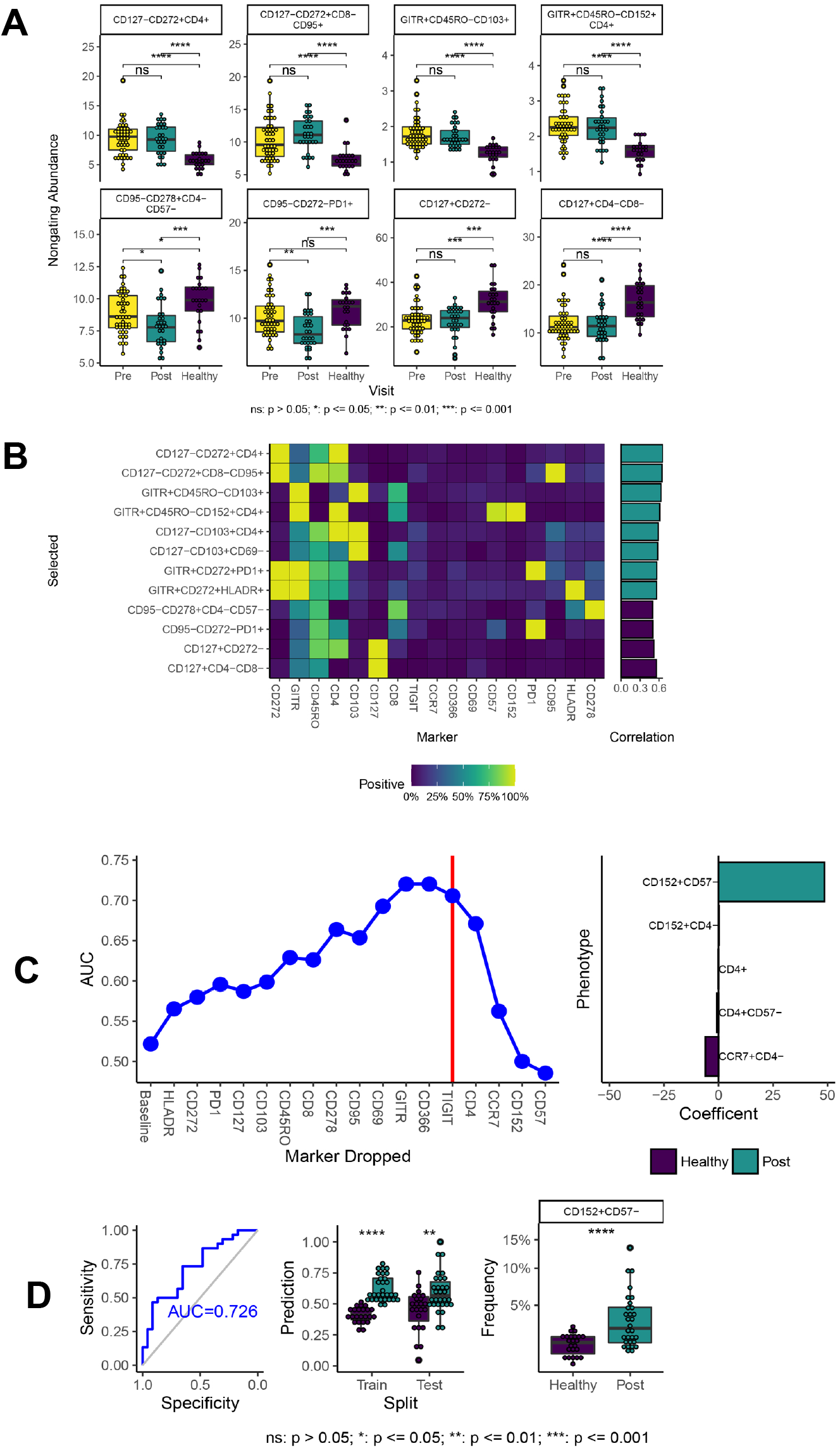
A) Distributions of the most statistically significant phenotypes across patient groups (post-treatment cHL patients vs. healthy donors; Checkpoint Panel; non-gating approach). B) Heat map depicting marker frequency (columns) within each phenotype. Adjacent bar graph shows correlation between population frequency and patient group. C) RFE analysis identifies CD4, CCR7, CD152, and CD57 as markers necessary to identify differences between patient groups. D) Model, validation, and data for selected RFE-defined phenotypes.

